# Thalamic connectivity topography in spina bifida newborns is linked to functional lesion level

**DOI:** 10.1101/2023.06.22.23291762

**Authors:** Hui Ji, Kelly Payette, Anna Speckert, Ruth Tuura, Patrice Grehten, Raimund Kottke, Nicole Ochseinbein-Kölble, Cornelia Hagmann, Luca Mazzone, Martin Meuli, Beth Padden, Annette Hackenberg, David-Alexander Wille, Ueli Moehrlen, Beatrice Latal, SPINA BIFIDA STUDY GROUP ZURICH, Andras Jakab

## Abstract

Spina bifida affects spinal cord and cerebral development, leading to motor and cognitive delay. We investigated whether there are associations between thalamocortical connectivity topography, neurological function and developmental outcomes in open spina bifida. Diffusion tensor MRI was used to assess thalamocortical connectivity in 44 newborns with open spina bifida who underwent prenatal surgical repair. We quantified the volume of clusters formed based on the strongest probabilistic connectivity to the frontal, parietal, and temporal cortex. Developmental outcomes were assessed using the Bayley III Scales, while the functional level of the lesion was assessed by neurological examination at two years of age. Higher functional level was associated with smaller thalamo-parietal, while lower functional level was associated with smaller thalamo-temporal connectivity clusters (Bonferroni-corrected p<0.05). Lower functional levels were associated with weaker thalamic temporal connectivity, particularly in the ventrolateral and ventral anterior nuclei. No associations were found between thalamocortical connectivity and developmental outcomes. Our findings suggest that altered thalamocortical circuitry development in open spina bifida may contribute to impaired lower extremity function, impacting motor function and independent ambulation. We hypothesize that the neurologic function might not merely be caused by the spinal cord lesion, but further impacted by the disruption of cerebral neuronal circuitry.

## Introduction

Spina bifida (SB) is a prevalent congenital disorder arising from the defective fusion of neural tube. Spinal bifida aperta (SBA) is the most common type of open neural tube defects, characterized by the exposure of the placode to the amniotic fluid. Individuals with spina bifida may experience a range of clinical complications, consisting of varying degrees of lower extremity motor dysfunction, neurogenic bladder and bowel dysfunction, Chiari II malformation, and an increased risk of cognitive and learning problems.^1,2^ The anatomical setting exposes the developing neural tissue during pregnancy to the damaging effects of amniotic fluid and mechanical trauma. In recent years, prenatal repair has been established to prevent further damage to the exposed spinal cord as well as to ameliorate hindbrain herniation associated with the Chiari II malformation and hydrocephalus.^3–5^

With fetal repair surgery, motor development and function improved compared to postnatal surgery.^5,6^ However, despite advances in surgical repair, most infants who followed prenatal or postnatal surgical repair still exhibit varying degrees of impaired lower-limb motor function, neurogenic bladder, and impaired bowel function.^7,8^ It is important to note the outcome can vary widely, and some cases show mild or no lower limb motor impairment and good bladder control. Additionally, individuals with spina bifida also experience a variety of developmental impairments, such as deficit in cognitive skills.^9^ However, the neural correlates of such impairments are not fully understood. The heterogeneity of symptoms in spinal bifida aperta necessitates the identification of pre- or early postnatal markers of developmental impairments and neurological deficits.

MRI studies provide evidence of structural brain abnormalities in individuals with SB. Structural MRI-based quantitative morphometry demonstrated a remarkable heterogeneity in brain development. Significant reductions in grey matter (GM), white matter (WM) and subcortical structure volumes were described in children with SB.^10–12^ In addition, surface-based analyses reported reduced cortex volume, disrupted cortical thickness and gyrification in SB.^10,11^ A recent study examining the morphology of fetuses following prenatal repair revealed significantly different brain volumes and gyri shape index compared to control fetuses.^13^

The thalamus is a central relay station of inputs to the cerebral cortex, and is involved in processing information. Thalamocortical (TC) connectivity, consisting of the complex network of fibers linking the thalamus and the cortex, is a fundamental contributor for the formation and the maintenance of cerebral connections essential for proper brain function. This intricate TC circuitry emerges rapidly during mid-gestational fetal development in humans. Altered TC connectivity has been found to be associated with later cognitive development in prematurely born infants.^14,15^ Therefore, to better distinguish markers of developmental impairments and neurological deficits, it is an appropriate and novel approach to examine neural structures, such as TC circuitry in spina bifida. However, the role of TC connectivity impairments in SBA is not yet understood. Diffusion MRI (dMRI) was used in several studies to investigate white matter^16–18^ and deep grey matter microstructure in SB.^19–21^ The abnormal diffusion indices in these structures may represent the microstructural basis for postnatally altered TC connectivity development, and provide a possibility of using dMRI based tractography between the thalamus and cortex to explain later impaired outcomes variability in SB.^21–23^ To the best of our knowledge, no analysis has been published on the structural TC connectivity in SBA newborns who underwent prenatal repair, and its possible relationship to postnatal impaired neurodevelopmental outcomes requires further evaluation.

We hypothesize that early damage to the rapidly developing TC circuitry may in part explain the later manifesting neurodevelopmental impairments, and we hypothesize that there is a link between the underlying spinal lesion, functional deficits and impaired development of TC circuitry. In addition, we hypothesize that the altered cerebrospinal fluid circulation as a consequence of impaired spinal cord development may exert damage on the developing mid-brain and diencephalic structures, leading to impairments in TC circuitry development. Therefore, the aim of our study was to examine these links in a cohort of newborns who underwent prenatal repair for SBA and who have been examined at two years of age.

## Methods

### Study population, clinical and neurodevelopmental variables

The infants in this study were drawn from a prospective cohort of infants who underwent open prenatal repair for spina bifida from June 2014 to June 2020. The original inclusion criteria and the criteria to undergo fetal repair surgery, as well as the characterization of the general study cohort are found in previous publications of the research team.^5^

In the present MRI study, the criteria for subject enrollment were the availability of written informed consent for the further use of data in research, the availability of good quality MRI (structural and DTI) performed at newborn age, availability of 2-year developmental outcomes and functional level assessments. All MRI data of the study were evaluated for quality by a researcher with 10 years’ experience in fetal and infant MRI. Cases were excluded if DTI was not available or if there were motion or other imaging artifacts that affected more than 10% of the DTI frames.

The functional level was assessed from newborn age through the corrected age of 24 months by pediatricians and neurologists or pediatric rehabilitation specialists at the Zurich Center for Spina Bifida, University Children’s Hospital Zurich and shown almost consistent results across multiple evaluations from early postnatal through 2 years of age. The functional level was determined by evaluation of muscle strength in the lower limb: extension and flexion of the hip, knee and ankle, hip abduction and adduction and inversion and eversion of the ankle. Functional level is defined by the lowest myotome with normal strength (M5) of the innervated muscles. In this study, partial innervation was defined by reduced muscle strength (M1-4) in more distal myotomes. Developmental assessments were based on the Bayley Scales of Infant and Toddler Development, Third Edition (Bayley-III), performed at 24 months of age by child development specialists at the Child Development Center, University Children’s Hospital Zurich. The Bayley–III assesses different developmental domains providing three composite scores: cognitive composite score (CCS), language composite score (LCS) and motor composite score (MCS). The evaluation was conducted by trained developmental paediatricians. Each of these has a mean score of 100 and a standard deviation of 15. Raw scores were transformed into standard scores using the American norms.^24^

Forty-four SBA newborns with mean (standard deviation) gestational age at birth of 35.5 (±1.8) weeks and at MRI of 37.97 (±1.13) weeks met these criteria and were enrolled in the analysis. The flow chart of how the final study population was reached is summarized in **Figure 1**. Further details of the patient demographics are found in **Table 1**.

**Table 1.**
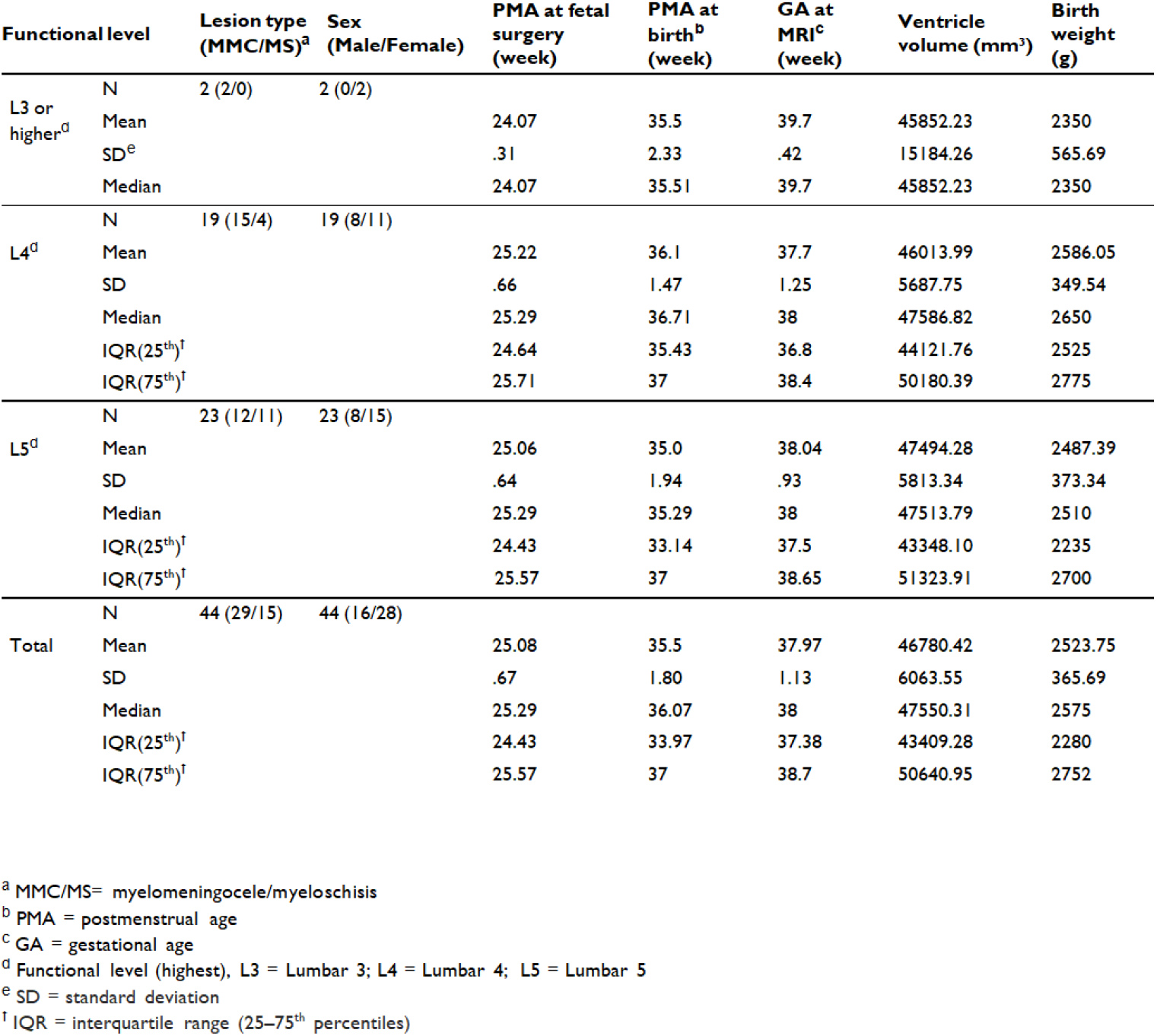
Patient demographics and comparison between groups based on the functional level at 2 years of age.

**Figure 1.**
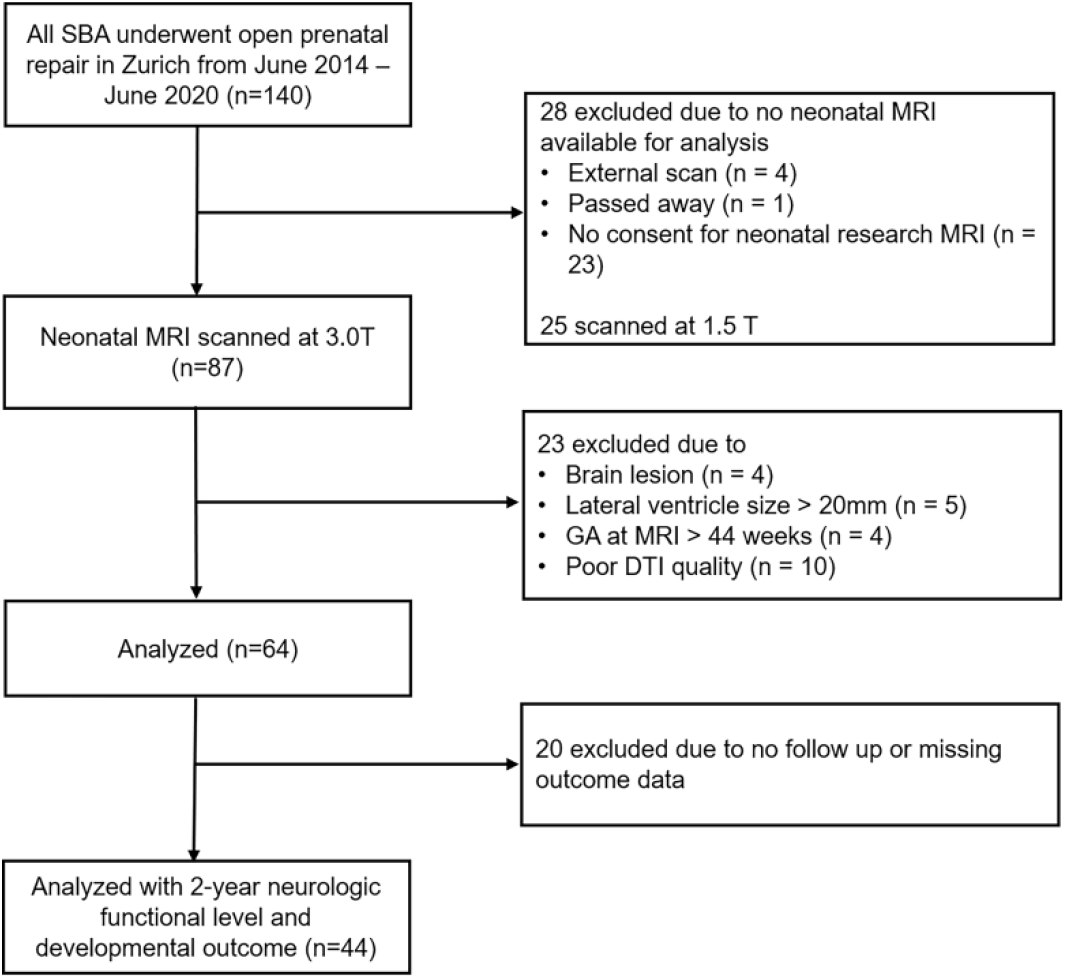
Flow chart of inclusion criteria applied to the study cohort.

All parents or caregivers gave written informed consent for the further use of their infants’ data in research. The ethical committee of the Canton of Zurich approved the studies for collecting and analyzing clinical data retrospectively (2016-01019, 2021-01101 and 2022-0115). The clinical variables in our work are based on a data registry that was established to collect all pertinent data in a prospective and systematical way. All the data used in this study were obtained from this REDCap™ based repository.

### MRI Acquisition

Newborn MRI was performed on a 3.0T MR750 scanner (GE Medical Systems), using an eight-channel receive-only head coil. All infants were sedated during scanning. Ear protection was used, oxygen saturation and heart rate were monitored, and all examinations were supervised by a neonatologist or a neonatal nurse. Structural, T2-weighted MRI was performed with a fast recovery fast spin echo sequence (FRFSE, image resolution = 0.7 × 0.7 × 1.5 mm^3^, repetition time (TR) = 5900 ms, echo time (TE) = 97 ms, flip angle: 90°, matrix: 512 * 320, slice thickness: 2.5 mm, slice gap: 0.2 mm) in axial, sagittal and coronal planes. In case the radiographers detected motion or other artifacts, these scans were repeated. Diffusion tensor imaging (DTI) was acquired using a pulsed gradient spin echo planar imaging sequence with TE/TR = 90/3950ms, field of view = 18 cm, matrix = 128 × 128, slice thickness = 3 mm. 35 gradient encoding directions with *b* = 700 s/mm^2^ and four *b* = 0 images were acquired.

### Structural MRI processing and spina bifida template construction

The structural MRI was assessed for brain lesions. A super-resolution slice-to-volume reconstruction algorithm^25^ was applied to the three orthogonal T2 images, creating a 3D super-resolution reconstructed T2 volume brain (further referred to as 3DT2 image) with an isotropic image resolution of 0.5 × 0.5 × 0.5 mm^3^.

The 3DT2 images were segmented into tissue classes according to the definition of the developing Human Connectome Project (dHCP) structural pipeline.^26^ We utilized an in-house network based on the U-Net architecture,^27^ which was trained on 3DT2 images and ground truth image labels sampled from a population of normally developing neonatal controls and SBA data. The ground truth annotations were created in two steps: (1) running the dHCP structural pipeline on the selected normal and SBA cases, and (2) performing manual corrections for cases with errors, particularly in the presence of ventricular dilatation. The network was trained in a semi-supervised way, with initial segmentation taken from running the subjects through the dHCP structural pipeline, and then underwent iterations of retraining and manual correction. The volumes of the ventricle system (lateral, third and fourth ventricles) were calculated from these automated segmentations. The final segmentations were checked visually.

Next, a custom SBA T2-weighted template and region-of-interest (ROI) system was created for this study. Initially, 29 selected subjects’ 3DT2 images was co-registered to a 38-week neonate template from the neonatal atlas by Gousias *et al*.^28^ by using 6 degrees of freedom registration. The neonatal template served only as a spatial reference for initial alignment and defining image dimensions. This age of the initial target template was chosen as the mean gestational age at MRI of our study cohort was 38.11 weeks. Cases with the best 3DT2 image quality were selected from the SBA cohort. Next, an unbiased non-linear template representative of the SBA population (further referred to as SBA template) was reconstructed using the Advanced Normalization Tools (ANTs)^29^ by running the template reconstruction script. To ensure that the subjects used for template creation represented the mean width of ventricular dilatation, we employed lateral ventricle size width measurements and excluded extreme case with large dilatation.

### Diffusion tensor MRI processing

The DTI data processing was performed using an in-house Bash script wrapping various, commonly used software libraries. The *eddy_cuda* command in the Functional Magnetic Imaging of the Brain Software Library (FSL)^30^ was used for slice-to-volume reconstruction to correct for eddy current and head motion-induced geometric distortions. Additionally, the *dtifit*^31^ routine in FSL was utilized for diffusion tensor and scalar maps estimation.

Spatial alignment of the diffusion space to the SBA template was carried out by registering the B_0_ image to the 3DT2 template using the subject’s 3DT2 image as an intermediate step. Bias field correction was performed on the B_0_ image using *N4ITKBiasFieldCorrection*^32^ in 3D Slicer. Next, the bias field corrected B_0_ images were aligned to the subject’s 3DT2 using linear registration *flirt*^33^ in FSL. Subsequently, the 3DT2 images were linearly and non-linearly registered to the SBA template image using antsRegistrationSyN script in the ANTs toolbox.^29^ The resulting linear transformation matrix was converted into an FSL compatible transformation matrix format, for which the *c3d_affine_tool* was used in c3d software package. We then concatenated the two linear transformation matrices (B_0_ to 3DT2 and 3DT2 to SBA template) into one matrix. The subjects’ 3DT2 image in the SBA template space was used for manually annotating the thalamus masks. To acquire a nonlinear deformation field compatible with the FSL probabilistic tractography algorithm, B_0_ images were linearly and non-linearly registered with corresponding 3DT2 in SBA template space using *fnirt*^34^ in FSL. Based on the standard recommendation of the developers of the probabilistic tractography algorithm in FSL, the linear and non-linear transformations were used to propagate masks and fiber tracking results from the SBA template space back to individual diffusion space, while performing the calculations in the native diffusion space, but interpolating the results in a high resolution template space.

### Connectivity based thalamus parcellation

We used the BedpostX^35^ method in FSL to estimate fiber orientations and their uncertainties. Connectivity-based thalamus parcellation was performed using the standard probabilistic tractography approach^36^ and cortical target-based clustering approach in FSL.^37^ The seed masks were manually annotated on each subject’s 3DT2 image in SBA template space for left and right thalami. Probabilistic tractography (PT) was performed for each voxel within thalamus mask, seeding from thalamus mask (5000 streamlines per-voxel) and to four target cortex masks. PT was performed separately for left and right thalamus, and an exclusion mask consisted of cerebral-spinal fluid (CSF) spaces, based on the segmentation of the 3DT2 image. For each subject, thalamus was segmented into four clusters based on seed-to-target connectivity pattern obtained from probabilistic tractography with *find_the_biggest* hard segmentation method in FSL^37^ by assigning each voxel within thalamus to the class with the highest connectivity probability.

### Statistical Analysis

TC connectivity was analyzed using two approaches: (1) volumetric analysis on the clusters obtained from the connectivity-based parcellation of TC connection. These clusters corresponded to projections from the thalamus to the frontal, parietal, and temporal regions and (2) voxel-wise analysis of the seed-to-target image maps which represent the voxel-level connection probability with the frontal, parietal, temporal and occipital cortices.

For the volumetric data, multivariate linear regression models were used and evaluated with SPSS 24.0^38^ and statistical visualizations were created by using R.^39^ For the connectivity strength maps, we perform cluster-wise statistical analysis using the Randomise tool^40^ in FSL. We corrected for multiplicity using threshold-Free Cluster Enhancement (TFCE)^41^

The associations with outcomes were tested with multiple univariate models, where the predicted variables (dependent) were the 2-year CCS, LCS, MCS or 2-year functional level. The CCS, LCS, MCS scores were continuous variables, while the 2-year functional level was encoded into an ordinal variable representing 3 levels: level of lumbar 3 segment or higher (referred to as L3), level of lumbar 4 (L4), level of lumbar 5 segment or lower (L5). In these statistical tests, no data was missing and no data imputation was carried out.

To investigate the relationship between the developmental and functional level variables and the standard space volume of each connectivity-based cluster, multivariate linear regression models were constructed. Bonferroni correction was employed to correct for multiple comparisons. In these models included variables were gestational age at MRI, lesion type, and ventricular volume as covariates. If volumetric differences were found, we ran a cluster-wise analysis on the voxel-level connectivity strength maps to see if more local relationships between TC connectivity and the dependent variables are driving these results, and whether a spatial predilection exists within the relatively large frontal, parietal or temporal clusters within the thalamus.

## Results

### Thalamocortical connectivity-based cluster volumes

The volumes of the TC connectivity-based clusters were mean (SD) mm^3^ for frontal lobe (2047.24 (485.24) mm^3^, parietal lobe (664.46 (407.95) mm^3^, temporal lobe (524.81 (309.11) mm^3^, occipital lobe (161.13 (185.09) mm^3^. We found that in 3 subjects, the volume of the occipital cluster was zero. This means that there were no voxels in the thalamus where the thalamo-occipital connections were stronger than the thalamo-frontal, parietal or temporal connections. Similarly, the variability of thalamo-occipital cluster volumes in the remaining subjects was high. Based on considerations of data quality and missing data, further statistical comparisons were restricted to multiplicity of three (frontal, parietal and temporal clusters), and occipital cluster volumes were excluded from the analysis.

### Association between structural thalamocortical connectivity-based cluster volume, 2-year developmental outcome and functional level

In our subject group, developmental outcome (mean (SD)) was lower than the norm (100 ± 15) for motor outcome and language outcome (MCS 78.93 (12.41), LCS 91.80 (10.59), respectively), but not for cognitive outcome (CCS 99.18 (11.96)). We found that the variability of the developmental outcomes (2-year LCS, CCS, and MCS) were not explained by the volumes of the frontal, parietal or temporal connectivity-based thalamus (Bonferroni adjusted *P* > 0.05).

The variability of the 2-year functional level (L3, L4 or L5) was explained by two models. Our study revealed a significant positive association between the volume of the parietal cluster in neonates and the 2-year functional level, with increasing parietal cluster volume being linked to lower (more favorable) functional levels (L5) **(Figure 2)**.

**Figure 2.**
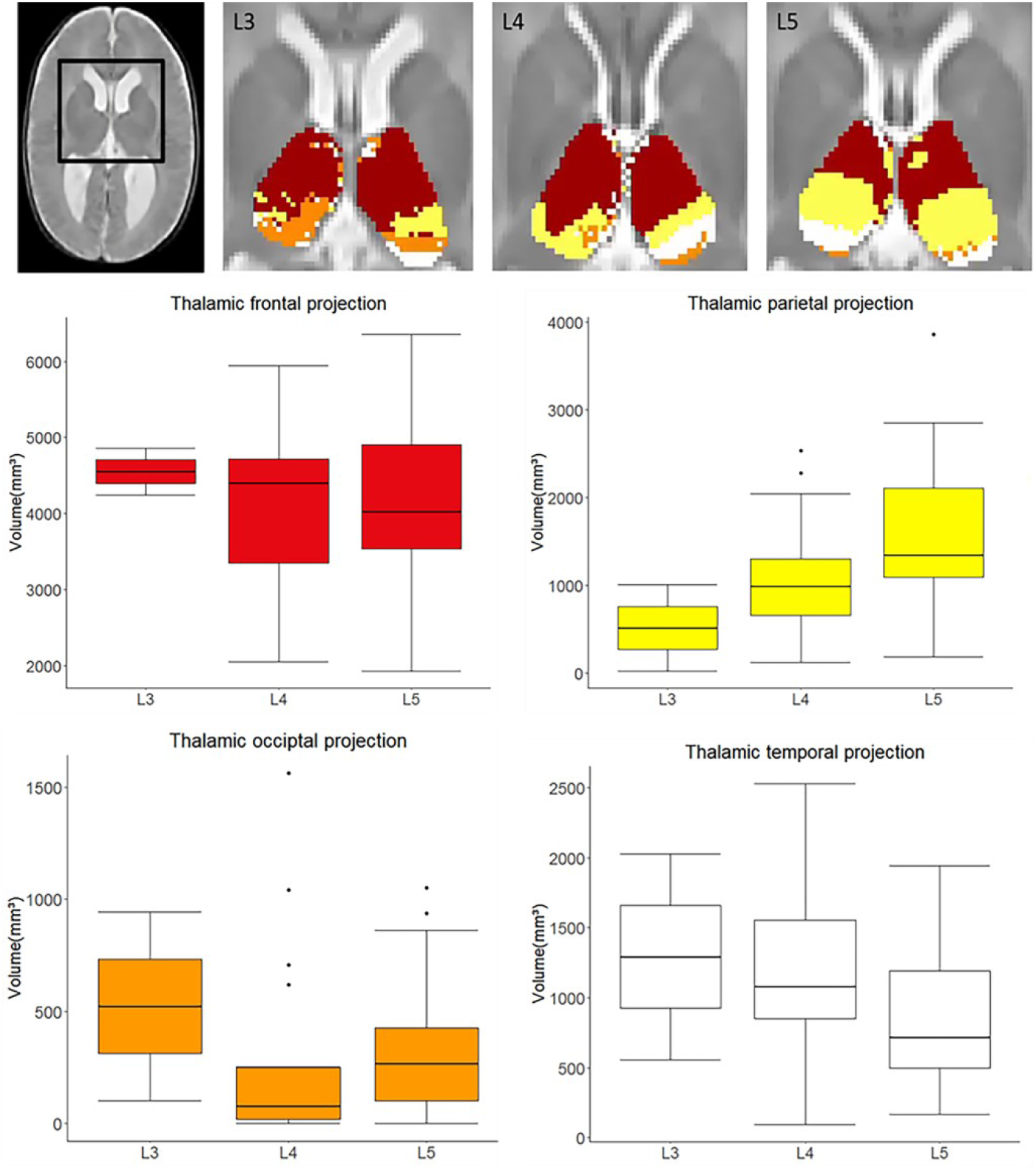
Thalamocortical connectivity-based parcellation is associated with the 2-year functional neurological level in newborns with spina bifida. Top row: in three selected cases, colored clusters represent the thalamocortical connectivity-based clusters in newborns, who had a functional level at L3, L4, L5, respectively. Results are displayed on top of the T2-weighted SBA template image. Middle and bottom rows: box-plots of each cluster volumes in three functional levels

**Figure 3.**
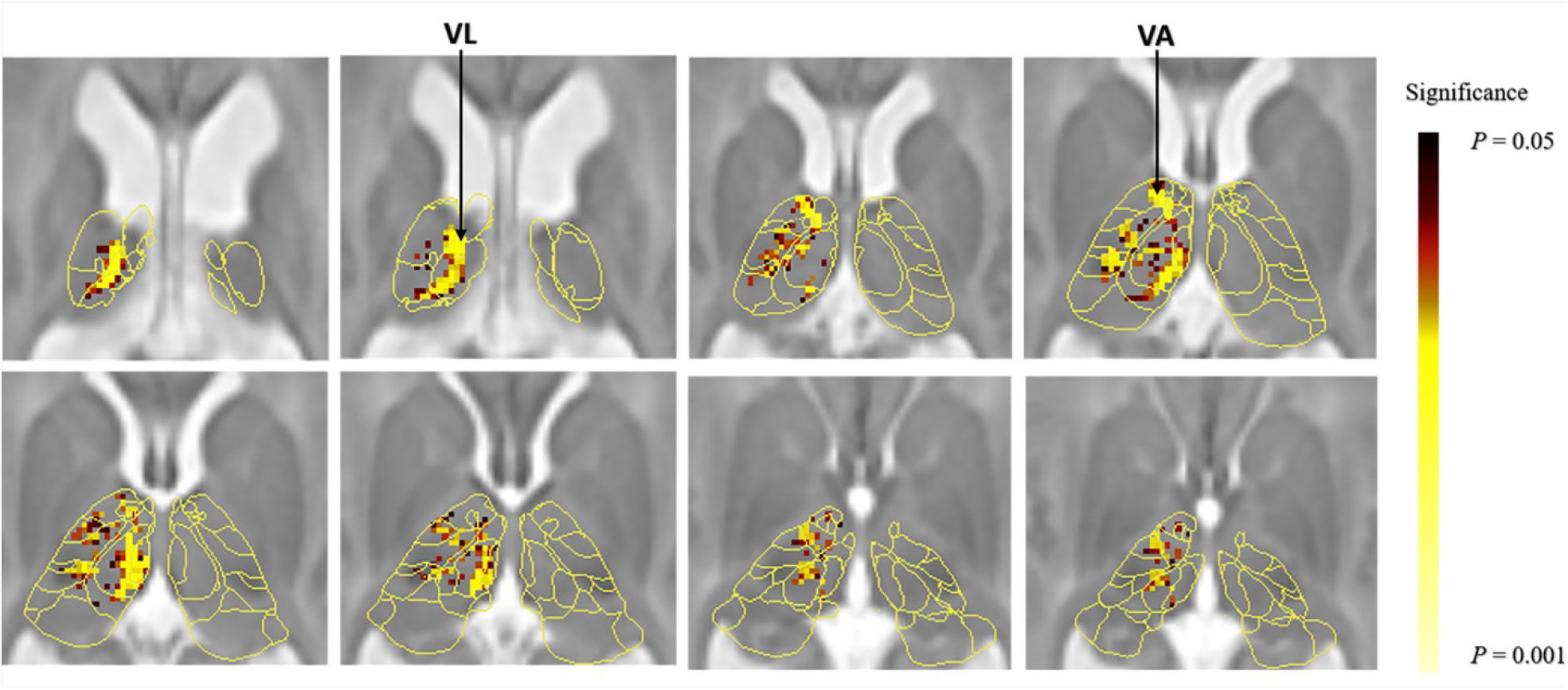
Voxel-wise analysis of the association between thalamocortical connectivity strength at newborn age and functional level at 2 years of age. Thalamic voxels where left thalamo-temporal connectivity was significantly (P < 0.05, TFCE-corrected) associated with functional level are shown as red and yellow overlay on a neonatal thalamus atlas and a T2-weighted SB template. VL, thalamic ventral lateral nuclei; VA, ventral anterior nuclei

A decreasing trend was observed for temporal cluster volume. No significant results were observed for the frontal cluster volume. The parietal connectivity-based thalamus cluster volumes (mean(SD) mm^3^ for level L3 (517.20 (692.92) mm^3^, level L4 (1086.59 (659.47) mm^3^, and level L5 (1599.70 ± 858.04) mm^3^ (**Supplementary Table 2**). The temporal connectivity-based thalamus cluster volumes were for level L3 (1295.51 (1038.92) mm^3^, level L4 (1258.48 (671.02) mm^3^, and and level L5 (855.98 (497.33) mm^3^ (**Supplementary Table 4**).

This first full model, which consisted of the parietal connectivity-based thalamus volume, gestational age at MRI, lesion type, and ventricular volume (term parietal cluster volume, *P* = 0.015, full model *P* = 0.013, Bonferroni adjusted *P* = 0.039), accounted for 19.6% of the variability in neurological function level (*adjusted R*^*2*^ = 0.196; **Table 2**). Among the terms in this model, lesion subtype (myelomeningocele vs. myeloschisis) was found to be significant, while ventricles volume or GA at MRI were not significant.

**Table 2.**
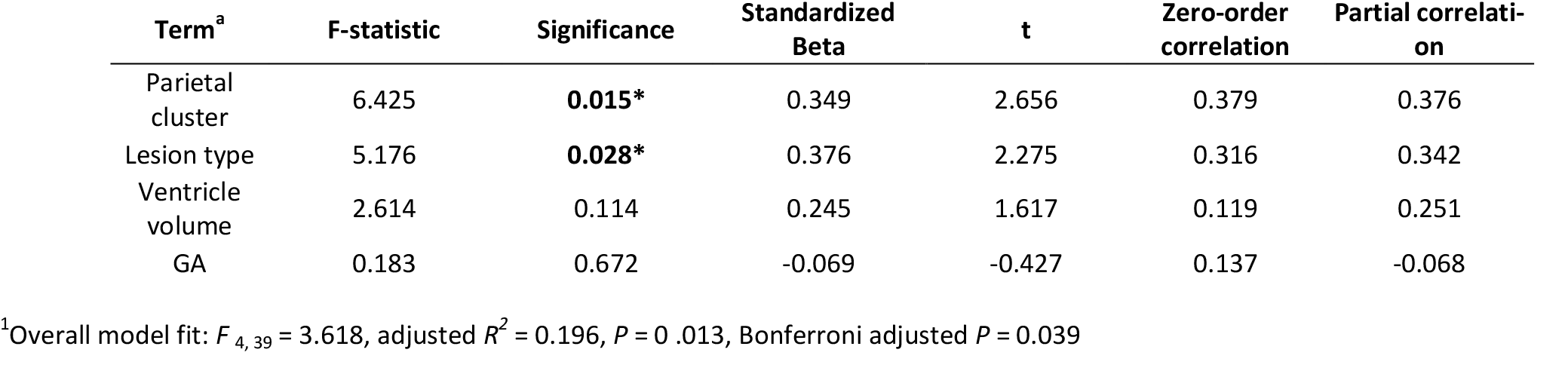
Results of multivariate linear regression analysis model 1.

| Term<sup>a</sup> | F-statistic | Significance | Standardized Beta | t | Zero-order correlation | Partial correlation |
| --- | --- | --- | --- | --- | --- | --- |
| Parietal cluster | 6.425 | **0.015\*** | 0.349 | 2.656 | 0.379 | 0.376 |
| Lesion type | 5.176 | **0.028\*** | 0.376 | 2.275 | 0.316 | 0.342 |
| Ventricle volume | 2.614 | 0.114 | 0.245 | 1.617 | 0.119 | 0.251 |
| GA | 0.183 | 0.672 | -0.069 | -0.427 | 0.137 | -0.068 |

<sup>1</sup>Overall model fit: $F_{4,39} = 3.618$ , adjusted $R^2 = 0.196$ , $P = 0.013$ , Bonferroni adjusted $P = 0.039$

A second multivariate linear regression full model was used to examine the relationship between neurological functional level at 2 years of age and the temporal connectivity-based thalamus cluster volume, gestational age at MRI, lesion subtype, and ventricular volume. However, after adjusting for multiple comparisons, this model was no longer significant (*P* = 0.032, Bonferroni adjusted *P* = 0.096). The analysis based on temporal connectivity-based cluster volumes revealed that 15.4% of the variance in neurological function level could be explained (*adjusted R*^*2*^ = 0.154; **Supplementary Table 1**). As with the first model, lesion type was a significant predictor, while neither ventricle volume nor GA at MRI contributed significantly to the model.

### Associations between voxel-level thalamocortical connectivity strength, developmental outcomes and functional level

A voxel-wise analysis revealed that the variability of the developmental variables (2-year CCS, LCS, and MCS) were not correlated with the thalamocortical connectivity strength (*P* > 0.05).

The same analysis on the TC connection strength and functional level revealed a significant negative correlation between functional level and the left thalamo-temporal connectivity (*P* = 0.014) (**Table 3**), indicating that lower functional level (L5) associated with reduced the thalamic-temporal connectivity. In contrast, while the left thalamo-parietal connectivity showed a non-significant trend toward positive correlation with functional level (*P* = 0.096) (**Table 3**), this suggests a potential relationship between lower functional level (L5) and increased thalamic-parietal connectivity

**Table 3.** Voxel-wise regression analysis of functional level with TC connectivity strength.

| Functional level | Left thalamo-parietal | Right thalamo-parietal | Left thalamo-temporal | Right thalamo-temporal |
| --- | --- | --- | --- | --- |
| L3 > L4 > L5 | 0.096 | 0.218 | **0.014** | 0.064 |

Peak p-values were shown corrected multiplicity with the TFCE method

## Discussion

To our knowledge, this is the first study evaluating structural thalamocortical connectivity and possible associations with neurologic function and developmental outcomes in SBA newborns. We found an association between the 2-year functional level and the topographical connectivity organization of the thalamus-parietal and thalamus-temporal cortex projections. However, no association with the 2-year developmental (reflecting the cognitive, language, and motor development) were found. The latter contradicts similar analyses conducted in preterm newborns, where thalamic circuitry was linked with later cognitive development.^14,15^ Similarly, the finding of a correlation between functional level and a lack of correlation with scores in the motor developmental domain presents a slight contradiction. This may stem from the fact that the neurological functional level assessment focuses mainly on lower extremity function in these cases (e.g. functional levels L3-L5), while the Bayley composite motor scores characterize motor development as a whole, including gross motor and fine motor functions of upper and lower extremities.

Our results align with those of previous studies that described altered MRI findings of reduced GM and WM volume,^12,42^ thinning cortex and enhanced cortical complexity in parietal and temporal region,^43^ and limbic tracts abnormalities,^22^ impaired parietal tectal cortical pathway,^21^ as well as altered DTI metrics of deep grey matter and white matter pathway that reveal the disrupted neurobiological process in SB.^17,44^ Similarly, an experimental animal study in a fetal sheep spina bifida model found reduced thalamic neuron numbers.^45^ Reduced WM volume, lower neuron numbers, thinning cortex or reduced GM volume in the parietal lobe might lead to a reduced projection in the thalamus, which in turn results in a changing topography of the TC connectivity-based clustering.

There is emerging evidence for a link between the development of neural circuitry and cognitive outcomes, however, many findings from the literature are ambiguous. Several previous studies on SB focused on brain morphology and diffusion metrics, and correlated these to developmental outcome. Wille *et al*.^46^ described no associations between impaired Bayley-III scores and overall brain structural abnormalities in the same cohort of SBA children after prenatal repair as presented in the current analysis. Treble *et al*.^11^ reported that cortical thickness and gyrification in SB was associated with IQ and fine motor dexterity assessed over a large time period between 8 and 28 years. Another study found diffusion properties of cortical-subcortical circuits correlated with executive function and fine motor performance in youth with SB.^47^ While no study so far has examined thalamocortical connections in SBA infants, two studies in preterm born infants, another population with altered brain development prior to term age, were able to show that structural thalamocortical connections could be a promising imaging biomarker for cognitive abilities at two years of age.^14,15^ In our study cohort, SBA infants who underwent prenatal repair surgery also showed a range of developmental and neurologic outcomes at two years of age, especially impairments of lower-extremity motor function (assessed with functional levels). However, we could not demonstrate a correlation between TC connections and cognitive outcomes at two years of age. However, the assessment of cognitive abilities is limited at two years of age and is based on the structured assessment of play behavior. It is possible, that an assessment at school or adolescent age may reveal a link between TC connections and cognitive, visuomotor and behavioral functions at school age as has been shown for school-aged preterm born children.^48^

The developmental timing of how TC connections emerge may explain our findings. SBA newborns were undergoing prenatal repair surgery at a mean gestational age of 25 weeks, which is still a very active time period of neural circuit development. After a rapid and early development in mid-gestation, thalamocortical circuits are mostly mature at the time of birth.^49,50^ Before the time of surgery, the first wave of neurons migrate to the subplate region and contribute to the formation of the first thalamus-subplate circuits, as well as shape the later maturing TC connections.^51,52^ This early organizational process may be vulnerable to disruptions from several factors, such as the presence of harmful molecules or osmotic forces from the amniotic fluid or the accumulation of cerebral spinal fluid that generate expansion forces.^53–55^ Meuli *et*.*al* described in fetal sheep model that the loss or degeneration of neural tissue may be attributed to the abnormal exposure of unprotected neuronal tissue to the chemicals from amniotic fluid during pregnancy.^56^ To explain some of findings, we assume that abnormal spinal cord development in spina bifida lead to abnormal (afferent sensory) inputs to the thalamus, thereby affecting the development of the thalamus and of the neural connectivity.^57,58^ In conclusion, while secondary damage to the developing central nervous system is ameliorated by fetal repair surgery by significantly reducing spinal cord exposure to amniotic fluid and mechanical damage, salvaging the early developing thalamocortical circuitry from damage may no longer be completely possible.

Our results showed a possible link between TC connection topography and functional level with the largest effect being between L4 and L5. To interpret this finding, it is important to understand the characteristics of the functional loss that is associated with these spinal levels. Several classification systems have been proposed to characterize and classify the functional deficits as a consequence of the spinal lesion in SBA.^59–61^ For example, the functional level could be described as the muscle strength corresponding to the muscle groups innervated by the spinal nerves of different anatomical levels. McDonald *et al*.^62^ proposed a classification system to predict SBA patients’ walking ability based on the assessment of muscle (iliopsoas) strength. In our study cohort, functional levels were at level range of lumbar 3 (L3) to lumbar 5 (L5). The most significant differences in TC connections were found between group L4 and group L5. According to the classification system employed in the Zurich Center for Spina Bifida, the differentiation of L4 and L5 functional level reflects differences in muscle strength of the legs, it is mainly made by assessing hip abduction, knee flexion and dorsal extension of the ankles. The examination of functional level in SBA allows for quantification of SBA functionality from early postnatal stages through age 2, yielding stable results across multiple assessments. This evaluation typically involves assessment of clinical history, functional observation, and muscle strength examination, which could be employed for counseling the prognosis of ambulation or walking ability. The TC connection differences between different functional levels in SBA neonates shed light on the fact that the overall neurologic function might not merely be caused by the anatomical level of the spinal cord lesion, but sustained or further impacted by disruption of cerebral neuronal circuitry. In SBA patients, this may further be exacerbated by hydrocephalus or hindbrain herniation.

Our findings demonstrate an altered thalamus connectivity topology in SBA neonates, likely caused by an increasing trend towards stronger thalamic-parietal connections in functional levels L3 to L5. Moreover, the analysis of TC projection (cluster) volume and functional level revealed a similar trend with TC connection strength. Specifically, lower functional levels (L5) were associated with more thalamic-parietal connections, while the opposite trend was observed for thalamic-temporal connections. Our findings suggest that these altered thalamus connectivity patterns may contribute to the functional deficit in SBA. These patterns are likely attributed to the TC connectivity parcellations defined by the cortical regions with the highest probability/connectivity within each thalamic voxel registered to a standardized template space. This way, the TC connectivity-based thalamus parcellation volumes in standard space have a competitive relationship with each other. Our findings suggest an asynchronous trend in thalamic connections with different cortical regions. Higher functional level (L3, or L4, more severe extremity motor deficit) exhibiting more severe disruption of thalamic parietal connectivity pathways. These findings are indirectly supported by Joannet *et al*.^63^ who reported a synchronized growth pattern of the thalamic substructures and the corresponding cortical connectivity. They also observed the prefrontal and temporal cortex thickness and their thalamocortical connections developed relatively faster than other brain regions within the first few weeks of life. Additionally, in neonates, the structural connectivity in the left hemisphere demonstrated higher efficiency compared to the right hemisphere. This may partially support our findings, as the connections in the left hemisphere thalamo-temporal region displayed a statistically significant correlation with functional level, while no significant correlation was observed in the right hemisphere.

Our study has the following limitations. Our results are limited by the relatively low case numbers (*n* = 44), which reduces the statistical power of our analysis. Furthermore, in the neonatal brain, axons are still in a pre-myelinated form in the majority of the white matter. The low diffusion anisotropy of non-myelinated white matter leads to higher uncertainty in estimating fiber orientations and connectivity. As a general limitation of diffusion MR based tractography, this method only traces relatively large fiber bundles, reducing the sensitivity in reconstructing the structural brain connectome. While we followed the TC connectivity analysis methodology previously reported, this method is still limited by the inherent noise in the DTI signal and the uncertainty in estimating the diffusion propagator or during the probabilistic tractography. The main limitation of the TC connectivity-based parcellation method is that it is a hard clustering method that is based on predefined cortical regions, which makes it less accurate in discriminating finer subdivisions of TC connectivity. Moreover, tractography in general is unable to resolve tract polarity, therefore TC connectivity in our manuscript (and in the majority of the DTI literature) refers to a mixture of thalamo-cortical as well as cortico-thalamic connections.

A further limitation is that our analysis excluded the occipital thalamic clusters, firstly based on missing data and data variability. The decision to restrict our analysis to these three clusters was also based on our hypothesis that functional level and developmental outcomes are more likely correlated with those TC connections that relay connections to cortices responsible for higher cognitive or motor functions, rather than visual connections. It is also important to note that the TC connectivity-based clusters are not independent observations, since they represent the subdivision of one structure into mutually exclusive compartments, so the relative volume change of one cluster affects the volume of the surrounding cluster(s).

The exact impact of spina bifida on neural circuit development depends on the type and location of the lesion, as well as the presence of other related conditions, such as hydrocephalus. Our results suggest that there are marked TC connectivity differences between the brains of newborns who will later have a functional neurological deficit that corresponds to L4 and L5 spinal levels.

In conclusion, our study highlights the potential clinical significance of differentiating between the L4 and L5 levels in individuals with spina bifida aperta. Our findings suggest that measuring TC connectivity could serve as a useful predictor for functional level and may aid clinicians in predicting the ability to walk independently. The novel findings presented in this study shed light on the potential clinical relevance of thalamocortical connectivity in spina bifida. The results of this study provide a foundation for future research aimed at investigating the potential impact of these findings on prenatal and postnatal diagnostics and prognosis. By further elucidating the neural mechanisms underlying spina bifida, we may be better equipped to develop more personalized and effective treatment strategies for children with this condition.

## Data Availability

The statstical data that support the findings of this study are available on reasonable request from the corresponding author. The original, patient-related, or raw/unprocessed imaging data are not publicly available due to patient confidentiality reasons.

## Acknowledgements

The authors want to first thank all families who participated in this research. In addition we thank our contributing study group without whom this research would not be possible. From the University Children’s Hospital this includes Barbara Casanova, Thomas Dreher, Ruth Etter, Patrice Grehten, Domenic Grisch, Annette Hackenberg, Cornelia Hagmann, Raimund Kottke, Niklaus Krayenbuehl, Claudia M. Kuzan-Fischer, Maya Horst Luethy, Markus A. Landolt, Andreas Meyer-Heim, Svea Muehlberg, Beth Padden, Silke Quanz, Brigitte Seliner, Mithula Shellvarajah, Sandra P. Toelle, Julia Velz, Alexandra Wattinger und Noemi Zweifel. From the University Hospital Zurich, our study group consists of Dirk Bassler, Lukas Kandler, Salome Meyer, Christian Schaer and Nele Struebing. Infrastructure support for this research was provided by the Clinical Trial Center, University Hospital of Zurich.

## Funding

This work has been supported by the URPP Adaptive Brain Circuits in Development and Learning (AdaBD) project, the Vontobel Foundation, the Anna Müller Grocholski Foundation, the EMDO Foundation, the Hasler Foundation, the OPO Foundation and the Prof. Dr. Max Cloetta Foundation. The University Children’s Hospital Zurich would also like to acknowledge a research contract with GE Healthcare

**This manuscript is the author’s original version**.

