## Supplementary Document for "Thalamic connectivity topography in spina bifida newborns is linked to functional lesion level"

### Part 1: Post-hoc statistical tests

The objective of the one-way ANOVA was to assess whether significant differences exist in temporal cluster volume among different neurologic functional levels (**Supplementary Table 1**). By comparing mean cluster volumes across functional levels, the ANOVA serve as an additional analysis to validate and reinforce the findings of the linear regression model, further supporting the association between neurologic functional level cluster volume.

To address the issue of imbalanced data and to capture the clinical significance of neurologic functional level, the three functional level groups (L3, L4, and L5) were regrouped into two categories: “Higher than or equal to L4” (L3/L4) and “Lower than L4” (L5). This grouping aimed to differentiate the functional levels that are clinically relevant for independent walking ability. Post-hoc pairwise comparisons were performed to assess specific differences between neurologic functional levels in the one-way ANOVA analysis.

The one-way ANOVA revealed a significant overall effect of neurologic functional level on thalamic parietal cluster volumes (p = 0.041, **Supplementary Table 2)**. Post-hoc pairwise comparison showed a significant difference between Group 1 and Group 2 (p = 0.018, **Supplementary Table 3).**

The similar one-way ANOVA was conducted to examine the relationship between neurologic functional level and temporal cluster volume. While the overall group comparison did not reach statistical significance **Supplementary Table 4**, post-hoc pairwise comparison revealed significant differences between Group 1 and Group 2 (p = 0.028, **Supplementary Table 5).**

**Supplementary Table 1.** Multivariate linear regression analysis of temporal cluster volume. The analysis account for confounding factors including GA at MRI, ventricles volume, and lesion type

| **Term** | **F-statistic** | **Significance** | **Standardized Beta** | **t** | **Zero-order correlation** | **Partial correlation** |
|---|---|---|---|---|---|---|
| Temporal cluster | 4.176 | **0.048** | -0.292 | -2.044 | -0.315 | -0.311 |
| Lesion type | 4.668 | **0.037** | 0.368 | 2.161 | 0.316 | 0.327 |
| Ventricles volume | 2.353 | 0.133 | 0.239 | 1.534 | 0.119 | 0.239 |
| GA | 0.03 | 0.863 | -0.029 | -0.174 | 0.137 | -0.028 |

[a]Overall model fit: $F_{4, 39}$ = 2.957, adjusted $R^2$ = 0 .154, $P$ = 0 .032, Bonferroni adjusted $P$ = 0.096

**Supplementary Table 2.** One-way ANOVA for thalamic parietal cluster volume and neurologic functional level.

| **Group** | **Functional level** | **N** | ***P*** | **Thalamic parietal volume (mm$^3$)** |
|---|---|---|---|---|
| 1 | L3 | 2 | | 517.26 ± 692.92 |
| 2 | L4 | 19 | **0.041** | 1086.59 ± 659.47 |
| 3 | L5 | 23 | | 1599.70 ± 858.04 |

**Supplementary Table 3.** Post-hoc pairwise comparison to assess specific differences between neurologic functional levels in the overall one-way ANOVA analysis.

| **Group** | **Functional level** | **N** | ***P*** | **Thalamic parietal volume (mm$^3$)** |
|---|---|---|---|---|
| 1 | L3/L4 | 21 | **0.018** | 1032.37 ± 666.89 |
| 2 | L5 | 23 | | 1599.70 ± 858.04 |

**Supplementary Table 4.** One-way ANOVA for thalamic temporal cluster volume and neurologic functional level

| Group | Functional level | N | *P* | Thalamic temporal volume (mm³) |
|---|---|---|---|---|
| 1 | L3 | 2 | | 1295.51 ± 1038.92 |
| 2 | L4 | 19 | 0.092 | 1258.48 ± 671.02 |
| 3 | L5 | 23 | | 855.98 ± 497.33 |

**Supplementary Table 5.** Post-hoc pairwise comparison to assess specific differences between neurologic functional levels in the overall one-way ANOVA analysis

| Group | Functional level | N | *P* | Thalamic temporal volume (mm³) |
|---|---|---|---|---|
| 1 | L3/L4 | 21 | **0.028** | 1032.37 ± 666.89 |
| 2 | L5 | 23 | | 855.98 ± 497.33 |

**Part 2: illustration of the image analysis pipeline.**

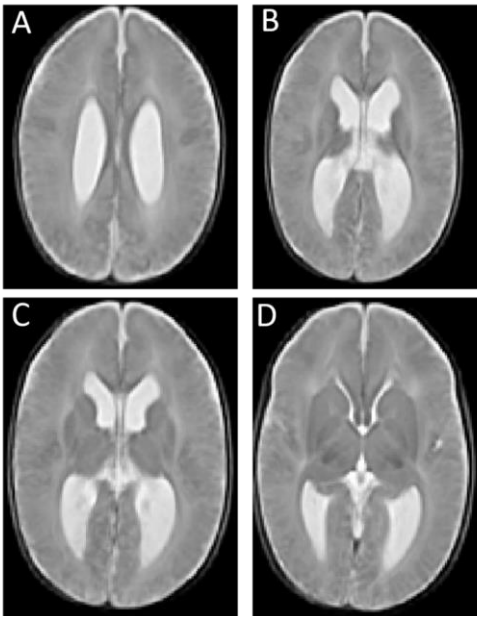


**Supplementary Figure 1**. T2-weighted template of the newborns of SBA. The images demonstrate the representative T2-weighted SBA template (A,B,C,D): with ventricular dilation (no visible T2 abnormality). The detailed methodology for creating the template is described in the methods section, where the process of selection, aligning/registration, and averaging the images to generate a SBA representative template is explained

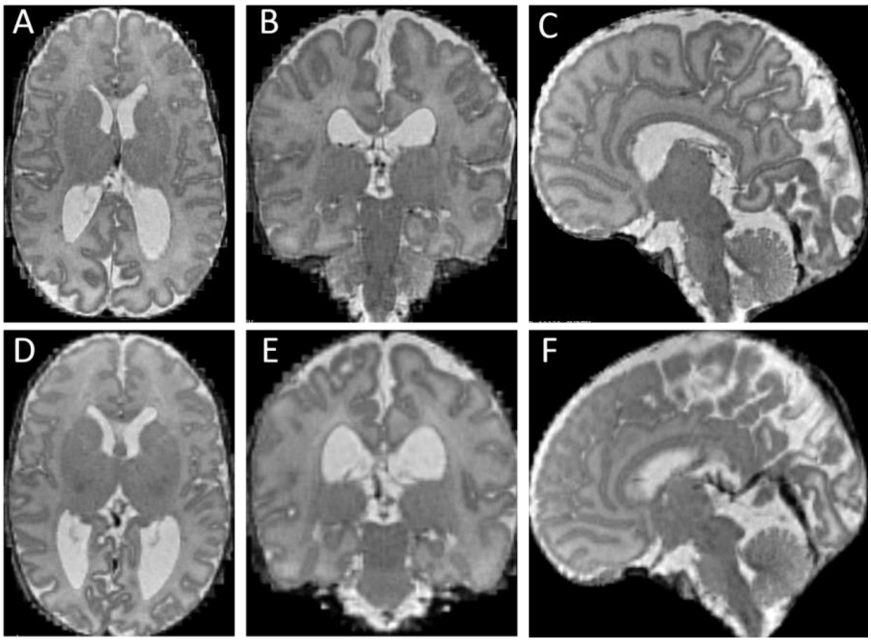


**Supplementary Figure 2**. An example of super-resolution reconstructed axial **(A**), coronal (**B**), and sagittal (**C**) plane T2-weighted images of a newborn with SBA. Axial (**D**), coronal (**E**), and sagittal (**F**) plane T2-weighted images of a SBA newborn registered to the SBA standard template space. The images demonstrate the successful alignment of the individual's anatomical structures to the standardized template, allowing for accurate comparison and analysis across subjects
